# Cooled Radiofrequency Ablation of the Genicular Nerves for Treatment of Chronic Knee Pain

**DOI:** 10.1101/2020.03.27.20045674

**Authors:** Yashar Eshraghi, Roshina Khan, Omar Said, Cruz Velasco, Maged Guirguis

## Abstract

**Background:** Chronic knee pain from conditions such as osteoarthritis (OA) is a significant problem in a growing and aging population. Cooled radiofrequency ablation (CRFA) is an emerging technique to treat chronic knee pain. There is significant literature noting the clinical outcomes of CRFA in anatomic locations including the peripheral joints and the lumbar spine. This retrospective study found significant improvements in Pain Disability Index (PDI) scores and Numerical Pain Rating Scale (NPRS) scores for patients with chronic knee pain who underwent cooled radiofrequency ablation (CRFA) therapy of the genicular nerves.

**Objectives:** This retrospective study evaluated the effectiveness of CRFA in the general chronic knee pain population.

**Study Design:** Retrospective electronic chart review.

**Setting:** Outpatient non-profit practice.

**Methods:** After institutional review board approval, we reviewed the data of 205 patients who had undergone cooled radiofrequency ablation therapy of the genicular nerves at a multiple-site pain practice between December 5, 2017 and September 4, 2019. This study’s primary outcome was improvement in Pain Disability Index (PDI) scores. The secondary outcomes were pain scores, assessed by the Numerical Pain Rating Scale (NPRS), and opioid consumption, assessed by daily Morphine Equivalent Dose (MED). From the 205 patients who met inclusion criteria, there were 104 patients who had PDI scores both before and after the CRFA procedure that were collected in the appropriate time frame. For these 104 patients, the pain scores and opioid consumption before and after the CRFA procedure were also collected. The age of the 104 patients ranged from 21 to 89 years. There were 38 males and 66 females.

**Results:** The mean PDI score before genicular nerve block and CRFA was 38.7, and the mean PDI score after CRFA was 26.5. After CRFA treatment, 67.38% of patients had a decrease in their PDI scores, 27.9% had no change, and 4.81% had an increase in their PDI scores. P-value <0.001 with 95% CI Median (-11, -7). The mean NPRS score before genicular nerve block and CRFA was 6.98, and the mean NPRS score after CRFA was 4.18. P-value <0.001 with 95% CI Median (-3, -2). The largest group of patients, 49% of patients, had a pain score reduction of 2.25 points, while the next largest group, 17.3% of patients, had a reduction of 0.75 points, followed by 12.5% of patients with a reduction of 3.75 points. When comparing Morphine Equivalent Dose (MED) before and after the CRFA procedure, 37.5% of patients were not on opioid medication at any time during the study; additionally, the MED did not change for the majority of patients (80.77%), while the MED decreased for 13.46% of patients and increased for 5.77% of patients. Mean MED before GNB and CRFA was 17.13 and 15.91 after CRFA. P=0.025 with 95% CI Median (0,0). No serious adverse events were reported.

**Limitations:** Retrospective nature of the study.

**Conclusions:** This study demonstrates the clinical effectiveness of CRFA for the treatment of chronic knee pain by improvements in PDI scores and NPRS scores for the majority of patients. Results from this study indicate that CRFA treatment provides significant pain relief and reduces the disability caused by chronic knee pain in a patient’s daily life.

## BACKGROUND

Chronic knee pain from conditions such as osteoarthritis (OA) is a significant problem in a growing and aging population. Mild and moderate OA can be treated with acetaminophen and non-steroidal anti-inflammatory drugs (NSAIDs). These medications, along with physical therapy and/or an exercise regimen, may help to maintain the patent’s mobility. Patients may also benefit from intra-articular knee injections. Corticosteroid injection provides significant short-term pain relief; however, multiple treatments put patients at risk for exacerbated cartilage destruction along with other side effects of steroid overuse (2). The injection of protein-based gel/fluids into the joint space to improve synovial fluid viscosity has shown moderate effectiveness; although this treatment hasn’t proven significant clinical utility (3). For patients with severe or late stage OA, a well-established terminal treatment is total knee joint replacement. Many patients may not be well-suited for this surgery due to issues of age, weight, smoking history, health, or a myriad of other factors. In addition, total knee replacement, while generally considered effective, has been associated in at least one study showing ongoing moderate to significant pain in up to 53% of patients (1).

Sensory innervation of the knee joint is accomplished by the articular branches of several nerves, known collectively as genicular nerves. The process of radiofrequency-generated lesions of the genicular nerves is referred to as genicular neurotomy. The standard for genicular neurotomy is thermal radiofrequency ablation, and there is literature to attest to its effectiveness for reducing knee pain. A prospective, randomized, double-blind controlled trial of genicular neurotomy in a series of 38 elderly patients who had severe knee pain due to OA and failed conservative therapy, showed statistically significant improvements in pain when treated with non-cooled radiofrequency ablation (4). Another prospective randomized study comparing radiofrequency neurotomy to diagnostic block alone showed 44% of the radiofrequency ablation treatment group rated “good or excellent” compared to only 12% of the block group (5). Both these publications address non-cooled radiofrequency ablation techniques.

Cooled radiofrequency ablation is a well-established method for delivering lesions into nervous tissue to reduce pain. There is literature noting the positive clinical outcomes of CRFA in anatomic locations including the sacroiliac joint and the lumbar facet joint (6, 7). Recently, there have been promising results from studies investigating the effects of CRFA when used for genicular neurotomy procedures to reduce knee pain from OA (8). There is room for further research exploring the effects and clinical outcomes of cooled radiofrequency ablation of the genicular nerves for treating chronic knee pain.

Previous studies using conventional, non-cooled radiofrequency ablation (RFA) demonstrated lasting and significant pain relief. However, the degree of pain relief from conventional RFA may be less than in those who received cooled RFA (9). The recent review by Kapural et al demonstrated significant improvement in pain score with no significant decrease of opioid use (10). Further investigation is indicated to show the long-term effects of cooled RFA and the potential superiority of cooled RFA therapy to conventional RFA therapy for treatment of chronic knee pain.

This retrospective review will follow the outcomes of cooled RFA over a long-term period to evaluate its effectiveness when used in patients with chronic knee pain by assessing patients’ pain disability index (PDI) scores before and after the CRFA procedure as a primary outcome. The PDI score is designed to measure the degree to which aspects of a patient’s life are disrupted by chronic pain and the overall impact of pain in a patient’s life. The PDI is a valid and reliable instrument to measure pain-related disability (11,12).

A secondary outcome was assessment of pain scores, as measured by the Numerical Pain Rating Scale (NPRS), which is a valid and verified measure of pain (13). Our next secondary outcome was opioid consumption, which was assessed by Morphine Equivalent Dose (MED).

## METHODS

After Ochsner Health System Institutional Review Board approval, we reviewed the charts of 205 patients who met inclusion criteria. Patients had to report at least a 50% improvement in pain after diagnostic genicular nerve block before proceeding with CRFA. Other inclusion criteria included age ≥ 18 years, chronic knee pain for longer than 3 months that interferes with functional activities, continued pain in the target knee despite at least 3 months of conservative treatments, PDI scores recorded 3 months before treatment with CRFA and at least 3 months after treatment with CRFA, radiologic confirmation of osteoarthritis for the index knee, and, for patients who had it, total knee arthroplasty completed without significant complications. Some of these patients were missing PDI scores either before or after the CRFA procedure, and they were excluded. Patients were excluded if they had evidence of inflammatory arthritis or other systemic inflammatory condition that could cause knee pain, previous or pending lower limb amputation, uncontrolled immunosuppression, chronic pain associated with significant psychosocial dysfunction, a clinically depressed state with Beck’s Depression Index score of > 22, history of substance addiction and abuse, and any patients who were pregnant during the study period. From the 205 patients we reviewed, we gathered 104 patients who had PDI scores both before and after CRFA treatment. The initial PDI scores, pain scores, and MED were taken before patients received genicular nerve blocks. The after PDI scores, pain scores, and daily MED were taken at least 3 months or later after the CRFA procedure. Data collected included patient identifiers, age, gender, diagnosis related to CRFA procedure, procedure dates, and dates when various scores were recorded.

Participants were aged 18 to 89 years old with a medical history of moderate to severe chronic knee pain for at least 3 months refractory to conservative therapy. Participants were patients treated at Ochsner Health System, Pain Management Clinic for knee osteoarthritis and chronic knee pain. Among the 104 patients, there were 38 males and 66 females.

During the CRFA procedure, skin anesthesia was achieved using lidocaine 1% over the respective injection sites. A 17 gauge 50 mm (4mm active tip) tip RF needle was slowly inserted and advanced to the junction of the lateral femoral and the epicondyle while contacting periosteum to block the superior lateral genicular nerve using the AP and lateral fluoroscopic imaging. The same process was repeated at the junction of the medial femoral shaft and the epicondyle to block the superior medial genicular nerve. The same process was repeated again using a 3rd needle, which was advanced at the junction of the medial tibial plateau and the epicondyle to block the inferior medial genicular nerve. The position of all needles was confirmed using AP fluoroscopy. Afterwards, lateral fluoroscopic images were obtained, and the needles were adjusted to 50% across the diaphysis. Motor stimulation at 2Hz up to 1.5V did not cause any radicular symptoms at any level. Each level was anesthetized with 1.5 cc of lidocaine 1%. Radiofrequency lesioning was performed for 150 seconds at 60 degrees. A combination of 3 cc of bupivacaine 0.25% with steroid was injected at all needle locations to block all targeted nerves.

## STATISTICAL ANALYSIS

Time between visits and PDI scores at each visit were characterized by minimum, median, maximum, mean and standard deviation. Change in PDI scores were subjected to tests for normality (Shapiro-Wilk, Anderson-Darling); which failed (p<0.03 for both). Thus, change in PDI score required non-parametric analyses: Wilcoxon signed rank test and distribution-free estimation of confidence intervals for median change in PDI scores. Goodness-of-fit tests aimed for an alpha>0.2, the Wilcoxon signed rank test alpha was set at 0.05, and the distribution-free confidence interval at 95%. Analyses were carried out in SAS 9.4.

Time between visits, Pain Scores (PS), and Morphine Equivalent Doses (MED) at each visit were characterized by minimum, median, maximum, mean and standard deviation. Change in PS and MED were subjected to tests for normality (Shapiro-Wilk, Anderson-Darling); which failed (p<0.01 for both). Thus, change in PS and MED required nonparametric analyses: Wilcoxon signed rank test and distribution-free estimation of confidence intervals for median change in PS and MED. Goodness-of-fit tests aimed for an alpha>0.2, the Wilcoxon signed rank test alpha was set at 0.05, and the distribution-free confidence interval at 95%. Analyses were carried out in SAS 9.4.

## RESULTS

The mean pain disability index (PDI) score before genicular nerve block and CRFA was 38.7, and the mean PDI after CRFA was 26.5. After CRFA treatment, 67.38% of patients had a decrease in their PDI scores, 27.9% had no change, and 4.81% had an increase in their PDI scores. P-value <0.001 with 95% CI Median (-11, -7). Figure 1 shows PDI scores for N=104 patients.

**Fig. 1.**
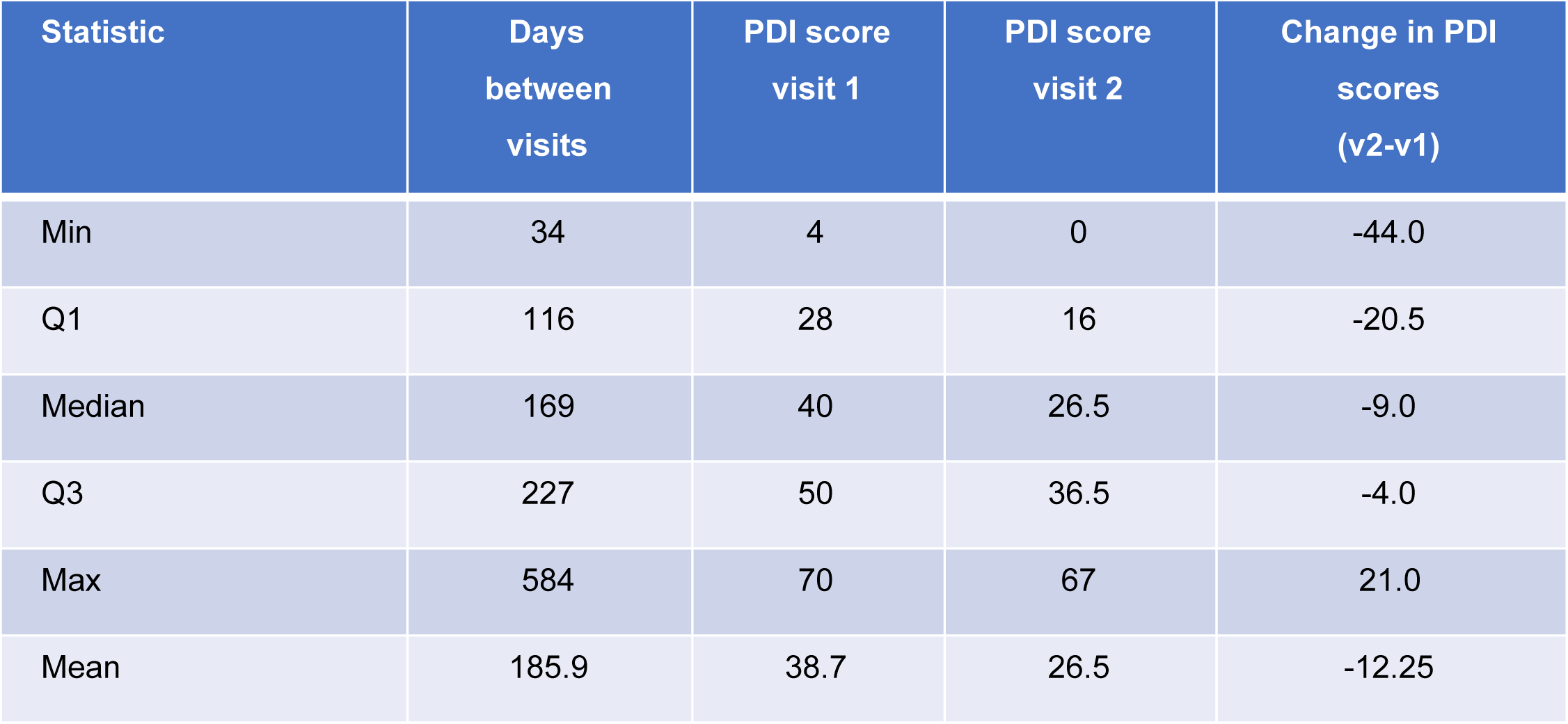
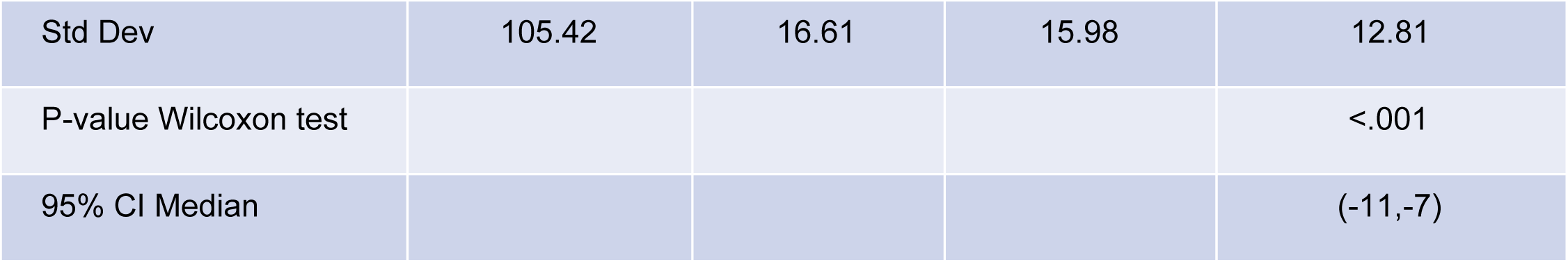
Comparison of PDI scores at visit 1, before CRFA procedure, and visit 2, after CRFA procedure.

Pain score refers to Numerical Pain Rating Scale (NPRS) score. The mean NPRS score before genicular nerve block and CRFA was 6.98, and the mean NRPS score after CRFA was 4.18. P-value <0.001 with 95% CI Median (-3, -2). The largest group of patients, 49% of patients, had a pain score reduction of 2.25; while the next largest group, 17.3% of patients, had a reduction of 0.75 points; and 12.5% of patients with a reduction of 3.75 points. Figure 3 shows pain scores for N =104 patients.

**Fig. 2.**
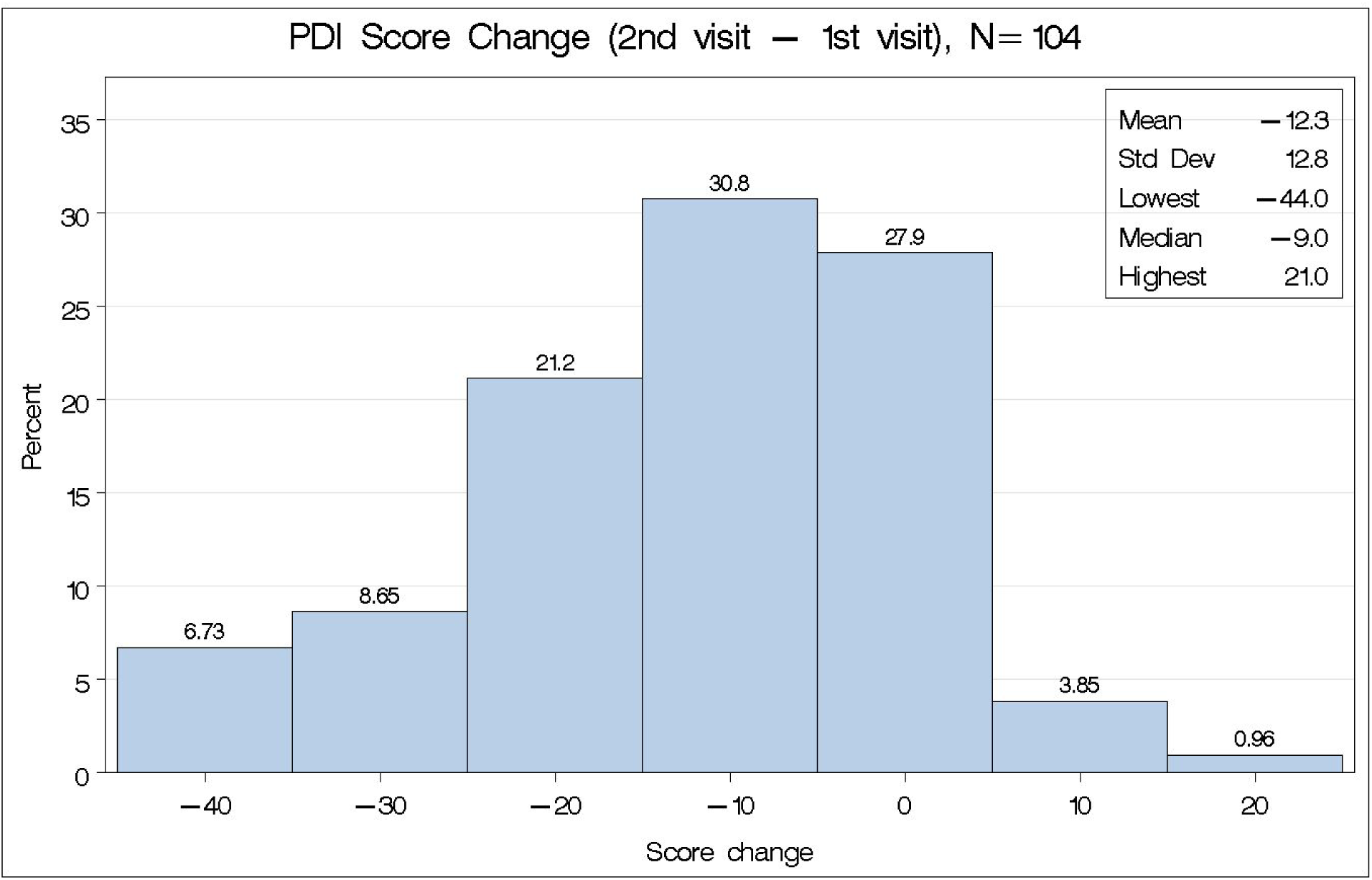
Percentage of patients whose PDI score changed by certain amounts between visit 1, before CRFA, and visit 2, after CRFA.

**Fig. 3.**
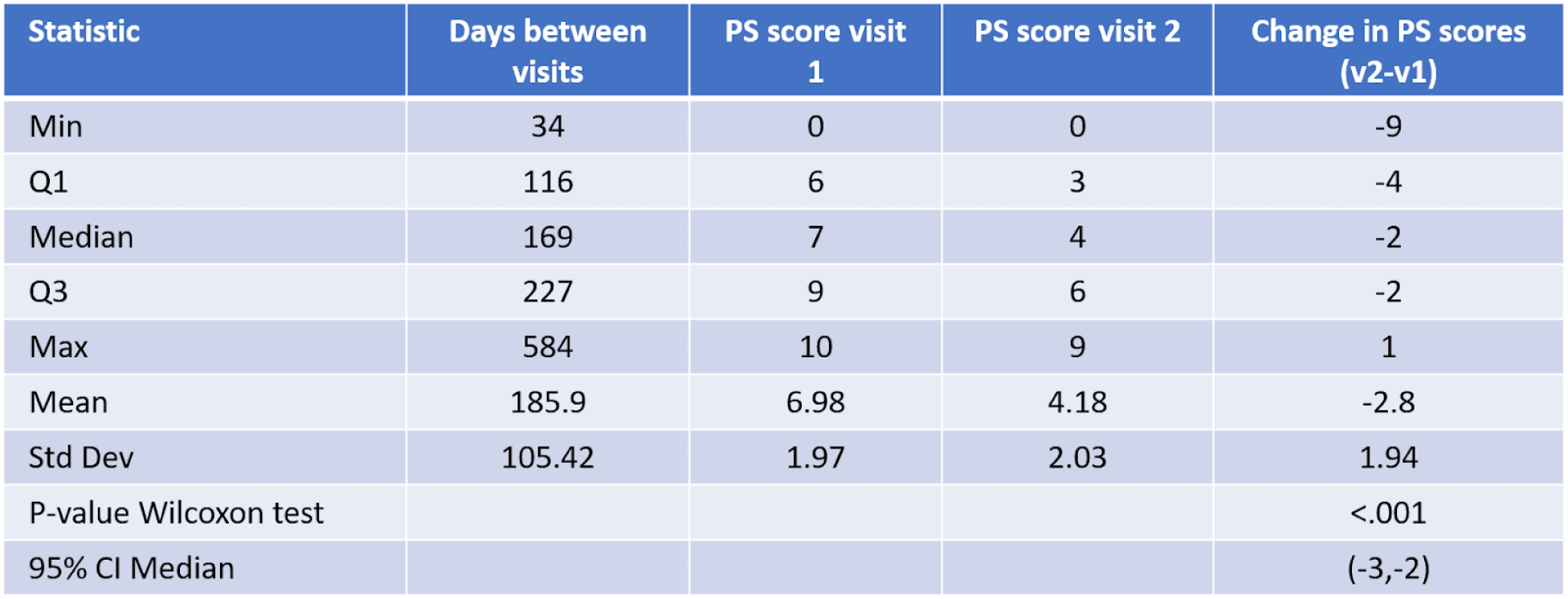
Comparison of pain scores at visit 1, before CRFA procedure, and visit 2, after CRFA procedure.

When comparing Morphine Equivalent Dose (MED) before and after the CRFA procedure, 37.5% of patients were not on opioid medication at any time during the study; and the MED did not change significantly for the majority of patients (80.77%), while the MED decreased for 13.46% of patients and increased for 5.77% of patients. Mean MED before GNB and CRFA was 17.13 and 15.91 after CRFA. P=0.025 with 95% CI Median (0,0). Figure 5 shows MED scores for N =104 patients.

**Fig. 4.**
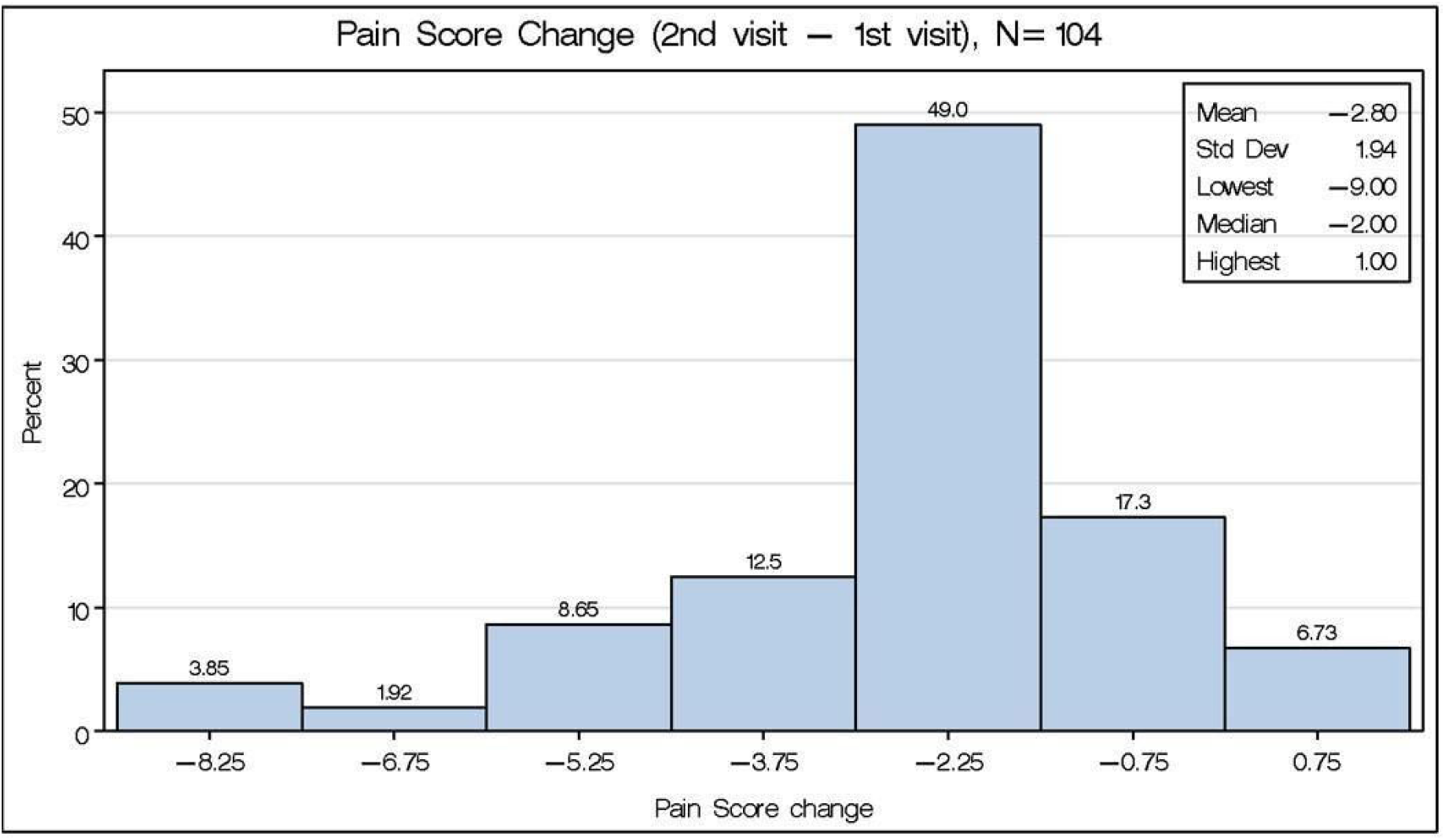
Percentage of patients whose pain score changed by certain amounts between visit 1, before CRFA, and visit 2, after CRFA.

**Fig. 5.**
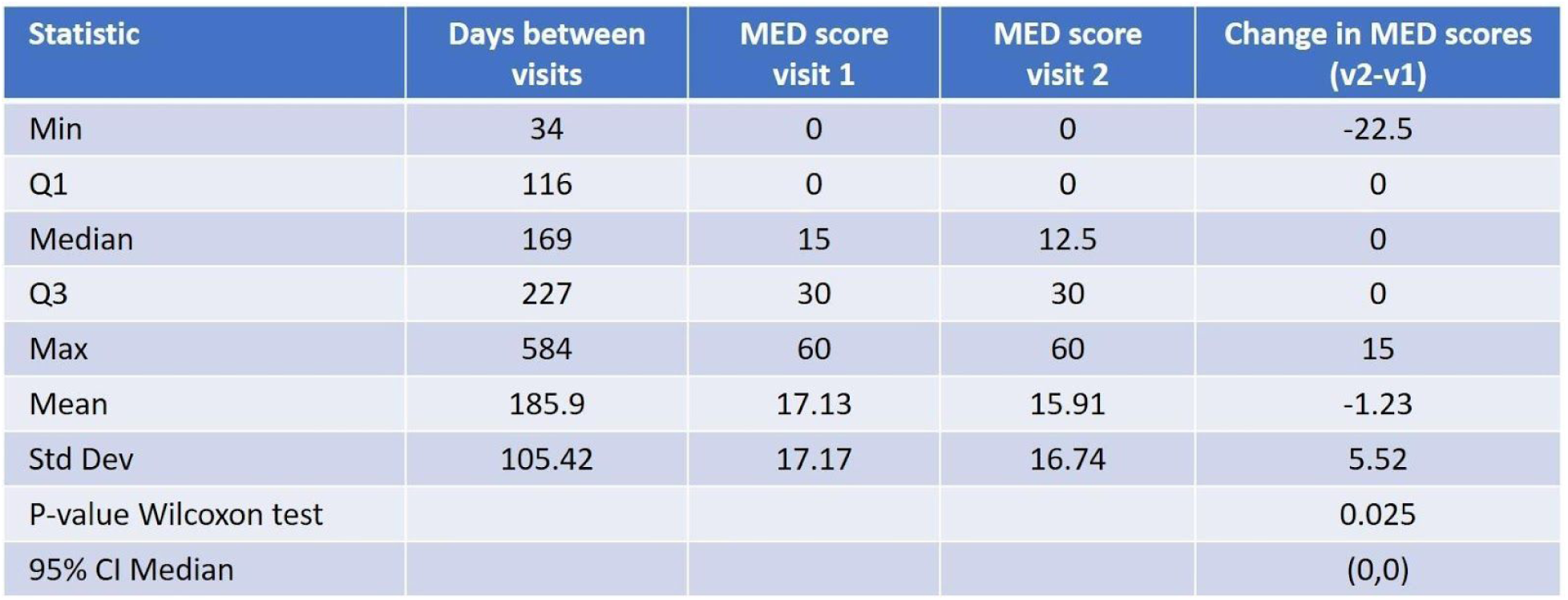
Comparison of MED scores at visit 1, before CRFA procedure, and visit 2, after CRFA procedure.

**Fig. 6.**
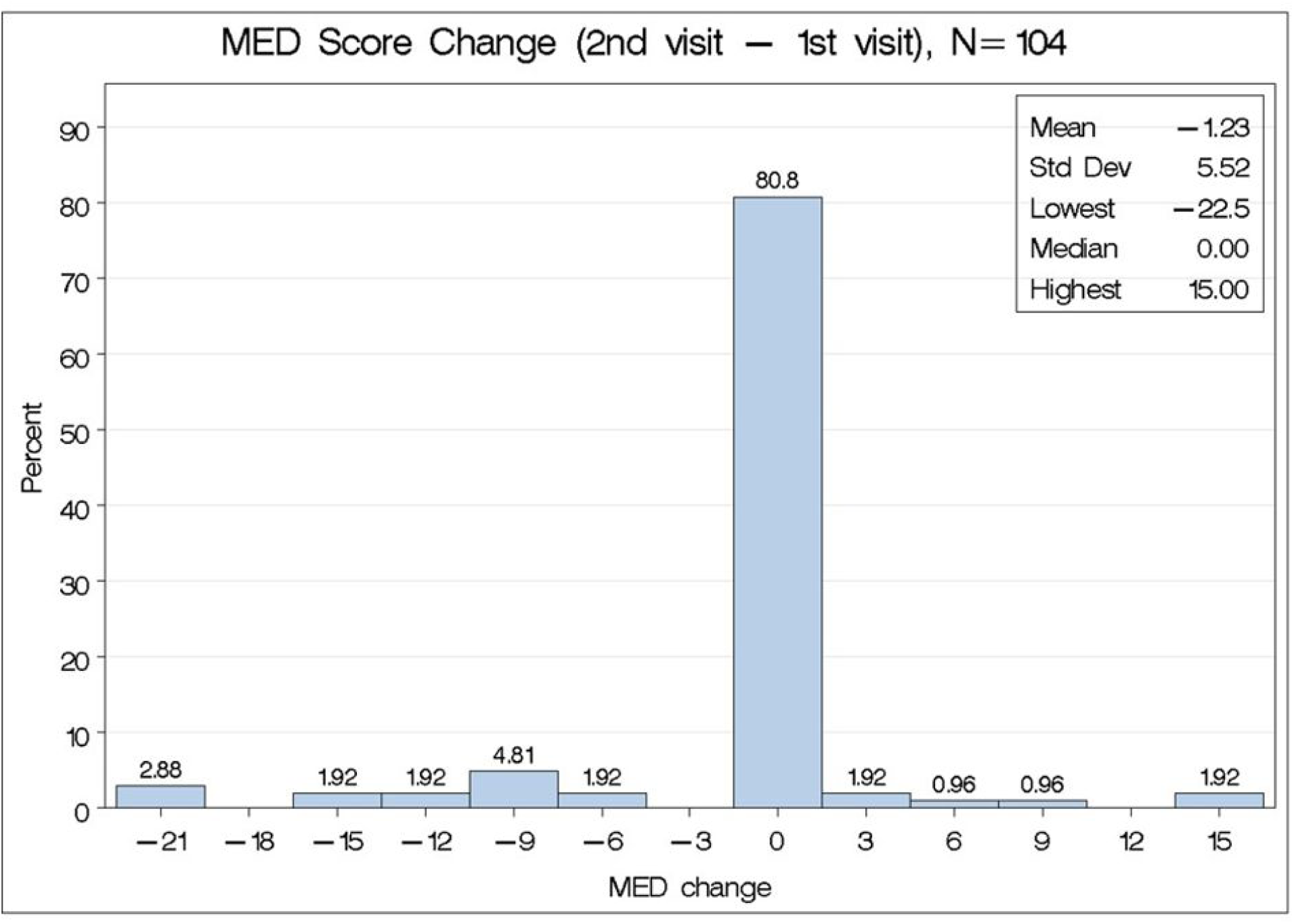
Percentage of patients whose MED score changed by certain amounts between visit 1, before CRFA, and visit 2, after CRFA.

No serious adverse events were reported.

## DISCUSSION

Cooled radiofrequency ablation is a safe treatment modality that can be performed to treat a variety of conditions. This particular study reviews the clinical efficacy of CRFA in treating chronic knee pain secondary to osteoarthritis. Functionality, reported pain scores, and change in morphine equivalent dose were all evaluated. This study provides evidence for significant improvements in pain disability index scores as well as reduction in patient reported pain scores. There was no significant reduction in the patients’ morphine equivalent dose.

The PDI score is designed to measure the degree to which aspects of a patient’s life are disrupted by chronic pain and the overall impact of pain in a patient’s life. Improvements in PDI scores indicate an improvement in quality of life for the patients in our review. The majority of patients in this study (67.38%) had a decrease in their PDI scores post CRFA treatment. The mean PDI score was reduced by approximately 31.5%.However, 32.7% of patients had either no change or an increase in PDI scores. It is worth noting that many of our patients had other sources of chronic pain besides knee pain contributing to their PDI score. This may explain why this group of patients did not have a reduction in PDI.

With respect to reported pain score, the majority of patients described a reduction in their overall pain levels. The largest group, approximately 50% of patients, had a pain score reduction of 2.25 points on the NPRS and approximately 93% of patients had some sort of pain relief following CRFA. This is an encouraging assessment of our measured secondary outcome.

Despite a majority of patients having a reduction in pain and an improvement in functionality, opioid consumption did not change for most patients. Only 13.5% of patients had a decrease in MED after CRFA. Approximately 81% of patients had no significant change in MED score and 5.8% actually had an increase in MED. Mean MED before genicular nerve block and CRFA was 17.13 and after CRFA mean MED was 15.91; however, this finding was not statistically significant [P=0.025 with 95% CI Median (0,0)]. There are a variety of potential explanations for the failure to reduce opioids. First, nearly 40% of patients were not on opioid medications before or during the study altogether. Additionally, as previously stated, many patients had other sources or comorbidities for which they may be taking opioids. Though it may be difficult, it would certainly be beneficial to investigate CRFA as a treatment option for patients who are taking opioids solely for chronic knee pain.

There were some limitations to this study. We were only able to gather PDI scores for 104 out of the 205 patients demonstrating a lost opportunity to review more robust data. To verify results, further research is warranted with a larger cohort comprising an even distribution of age groups and genders. Additionally, the post-CRFA PDI scores, pain scores, and MED were taken between 3 and 6 months after the CRFA procedure. We did not have a discrete point in time for follow-up that was consistent amongst our patients. Despite having some data on patients 12 months post-procedure, we were unable to consistently track a majority of the patients long-term through their post-procedure period. Being able to track more patients at least 12 months or more post-procedure would have enabled us to evaluate more closely the length of time patients may expect to have pain relief after CRFA. It would have also allowed us to assess whether pain and functional impairment returned suddenly or gradually over a period of time. Other limitations included the retrospective nature of the review and possible variability of technique in a tertiary pain practice with multiple pain providers.

This retrospective analysis confirms the results of previous trials on effectiveness of genicular cooled radiofrequency ablation to provide long term relief and improvement in quality of life for patients with chronic knee pain. Our results are comparable with Kapural et al report in terms of pain relief and opioid consumption. In addition, our review shows significant improvement in PDI scores in patients who received genicular nerve cooled radiofrequency ablation. Our study was able to demonstrate reproducibility of previous controlled trials in real-life practice patients, who likely had more than one pain source, so ultimately the effectiveness of the CRFA procedure relied on the care provided by multiple physicians.

Further research would be beneficial to compare CRFA therapy to conventional RFA therapy for the treatment of chronic knee pain. Though we did not have any serious adverse effects, it is also necessary to analyze the long-term effects of CRFA. It would be beneficial to further examine the effects of repeat CRFA as a treatment option for arthritic knee pain that has returned after previous CRFA. In previous studies regarding medial branch rhizotomy, repeat neuroablative procedures have been shown to be successful in restoring pain relief. Therefore, if knee pain does return, repeating the CRFA procedure seems to be a reasonable treatment option.

## CONCLUSIONS

The majority of patients in our study noted improvements in their pain disability index scores and pain scores post cooled radiofrequency ablation treatment. Results from this study indicate that cooled radiofrequency ablation treatment provides significant pain relief and reduces the disability caused by chronic knee pain.

## Data Availability

Data is available upon reasonable request.

## REFERENCES

1. Liu SS, Buvanendran A, Rathmell JP, et al. A cross-sectional survey on prevalence and risk factors for persistent postsurgical pain 1 year after total hip and knee replacement. Reg Anesth Pain Med. 2012 Jul-Aug; 37(4):415–22.

2. Arroll B, Goodyear-Smith F. Corticosteroid injections for osteoarthritis of the knee: meta-analysis. BMJ. 2004 Apr 10; 328(7444):869. Epub 2004 Mar 23.

3. Conrozier T, Jerosch J, Beks P, et al. Prospective, multi-centre, randomized evaluation of the safety and efficacy of five dosing regimens of viscosupplementation with hylan GF-20 in patients with symptomatic tibio-femoral osteoarthritis: a pilot study. Arch Orthop Trauma Surg 2009; 129:417–423.

4. Choi WJ, Hwang SJ, et al. Radiofrequency treatment relieves chronic knee osteoarthritis pain: A double-blind randomized controlled trial. PAIN 152 (2011) 481–487.

5. Ikeuchi M, Ushida T, et al. Percutaneous Radiofrequency Treatment for Refractory Anteromedial Pain of Osteoarthritic Knees. Pain Medicine 2011; 12: 546–551.

6. Stelzer W, Aiglesberger M, Stelzer D, et al. Use of Cooled Radiofrequency Lateral Branch Neurotomy for the Treatment of Sacroiliac Joint-Mediated Low Back Pain: A Large Case Series. Pain Medicine 2013 Jan: 14(1):29–35.

7. McCormick ZL, Walker J, Marshall B, McCarthy R, Walega DR. A Novel Modality for Facet Joint Denervation: Cooled Radiofrequency Ablation for Lumbar Facet Syndrome. A Case Series. Phys Med Rehabil Int. 2014;1(5):5.

8. Davis, et al, Prospective, Multicenter, Randomized, Crossover Clinical Trial Comparing the Safety and Effectiveness of Cooled Radiofrequency Ablation With Corticosteroid Injection in the Management of Knee Pain From Osteoarthritis. Reg Anesth Pain Med. 2017 Nov 1.

9. Bellini M, Barbieri M. Cooled radiofrequency system relieves chronic knee osteoarthritis pain: The first case-series. Anaesthesiol Intensive Ther 2015; 47:30–33.

10. Kapural, Leonardo et al.. “Long-Term Retrospective Assessment of Clinical Efficacy of Radiofrequency Ablation of the Knee Using a Cooled Radiofrequency System.” Pain physician 22 5 (2019): 489–494.

11. Chibnall JT, Tait RC. The pain disability index: factor structure and normative data. Arch Phys Med Rehabil. 1994;75(10):1082–6.

12. Soer R, Koke AJ, Vroomen PC, Stegeman P, Smeets RJ, Coppes MH, Reneman MF. Extensive validation of the pain disability index in 3 groups of patients with musculoskeletal pain. Spine (Phila Pa 1976). 2013;38(9):E562–8.

13. Jensen MP, Karoly P. Self-report scales and procedures for assessing pain in adults. In: Turk DC, Melzack R, editors. Handbook of Pain Assessment. New York: Guilford Press; 2011. pp. 19–44.

